# Standardising Workforce Cost Estimates Across Australian Jurisdictions: Genomic Testing as a Use Case

**DOI:** 10.1101/2024.04.09.24305541

**Authors:** Dylan A Mordaunt

## Abstract

**Introduction:** Labour costs are a key driver of healthcare costs and a key component of economic evaluations in healthcare. We undertook the current study to collect information about workforce costs related to clinical genomic testing in Australia, identifying key components of pay scales and contracts, and incorporating these into a matrix to enable modelling of disaggregated costs.

**Methods:** We undertook a microcosting study of health workforce labour costs in Australia, from a health services perspective. We mapped the genomic testing processes, identifying the relevant workforce. Data was collected on the identified workforce from publicly available pay scales. Estimates were used to model the total cost from a public health services employer perspective, undertaking deterministic and probabilistic sensitivity analyses.

**Results:** We identified significant variability in the way in which pay scales and related conditions are both structured and the levels between jurisdictions. The total costs (2023-24 AUD $) ranged from 160,794 (113,848 - 233,350) for administrative staff to 703,206 (548,011 - 923,661) for pathology staff (full-time equivalent). Deterministic sensitivity analysis identified that the base salary accounts for the greatest source of uncertainty, from 24.8% (20.0% - 32.9%) for laboratory technicians to 53.6% (52.8% - 54.4%) for medical scientists.

**Conclusion:** Variations in remuneration levels and conditions between Australian jurisdictions account for considerable variation in the estimated cost of labour and may contribute significantly to the uncertainty of economic assessments of genomic testing and other labour-intensive health technologies. We outline an approach to standardise the collection and estimation of uncertainty for Australian health workforce costs and provide current estimates for labour costs.

## Online Short Key Question Summary

### What is known about the topic?

Workforce costs are known to be a significant contributor to the cost of health services and complex health technologies.

### What does this paper add?

This paper systematically quantifies the variation in workforce costs across five Australian states, demonstrating that these differences are substantial enough to influence cost-effectiveness outcomes for genomic testing services. By identifying key areas of cost variation—such as base salaries, superannuation contributions, and allowances—this paper highlights how jurisdictional variations can contribute to differential health service costs, potentially impacting the affordability and sustainability of health programmes and technologies, across states.

### What are the implications for practitioners?

We provide reproducible estimates of workforce costs for use by health economists and policymakers conducting economic evaluations. The framework presented in this paper can also be adapted by other professionals in health economics and health workforce planning to assess jurisdictional variations and their implications for resource allocation and service delivery.

## Introduction

Workforce costs are the key driver of public health services costs ^1–3^. Workforce costs are also an important part of the economic evaluation of health services and technologies ^4, 5^. Australia has several different jurisdictions for employing and remunerating health workers, with a large number of resulting policy instruments contributing to cost, such as awards and enterprise bargaining agreements. These jurisdictions include the states and territories, as well as the Australian Commonwealth. Australia also shares a health workforce market with New Zealand. The landscape of worker conditions is complex and can be challenging to assess within the country, let alone in comparison to other countries ^5, 6^. Differences in workforce costs are important in assessing cost variance between jurisdictions and countries, in health policy decisions and in understanding the impact of labour costs on economic evaluation ^7^.

Understanding state-level variation in the costs of Australia’s public health workforce is important for several reasons. Firstly, estimates of cost directly influence the allocation of resources and the feasibility of implementing health programs across different states. If there’s a significant disparity in costs, it raises questions about whether funding is sufficient and equitably distributed to support these initiatives effectively. Additionally, this variation affects the generalizability of economic evaluations, as cost-effectiveness in one state might not translate to another due to these differences ^8^. It’s important to differentiate whether these variations represent true differences in the cost of delivering services or if they stem from disparities in bargaining power, administrative efficiency, or other factors ^8^. Such distinctions are important for policymakers, as they could undermine the effectiveness of public health programs ^9^.

In the context of costing a complex health service and trying to understand the uncertainty of labour cost for this health service by jurisdiction ^10^, we identified a gap in available tools and resources for estimating public health workforce costs. A standardised Australian labour costing tool could be used to support comparative policy research within Australia ^6^ and with other countries ^5^. The application in this particular use case was to undertake a sensitivity analysis of clinical genomic testing by varying labour prices based on the estimated labour costs in Australian states ^10, 11^.

Our question was, what is the cost of labour in public clinical genomics laboratories in Australian states, how does this vary and what are the sources of variation? To answer this question, we set about collecting information about Australian public health pathology workforce costs to the service provider, modelling this information and estimating sources of uncertainty for use in a comparative health systems context. Our objective was to define a standardised model for workforce costs across relevant Australian jurisdictions that would allow us to understand the sources of variability within the estimated cost.

## Methods

This report follows the relevant sections related to costing studies, of the Consolidated Health Economic Evaluation Reporting Standards (CHEERS) checklist, 2022 revision ^12^.

### Study design

We undertook a microcosting study of health workforce labour costs in Australia. Microcosting is a method that involves disaggregating the component costs of a resource, into small and meaningful components ^13^.

### Data collection

Labour type was identified as part of a process mapping exercise undertaken in a micro-costing study of clinical genomic testing at the Victorian Clinical Genetics Service (VCGS) in Victoria, Australia ^11^. The labour type was then classified based on the position descriptions used in this jurisdiction. Job advertisements were identified on publicly accessible government and recruitment websites, and these were used to help identify the equivalent positions (by labour type) for each state and resulting pay scales and industrial instruments. Data was then collected on the workforce costs from publicly available industrial instruments, such as pay scales, including awards and enterprise bargaining agreements. These documents were evaluated for the components of remuneration and conditions, and sources of variation, such as the amount of annual leave included. Inputs were then extracted and populated into a spreadsheet. An archive of these documents is included as a supplementary archive file.

### Study Population and Perspective

The study population included states and territories for which there was a public genomics laboratory, at the time of the study (Victoria, South Australia, Western Australia, New South Wales and Queensland). The perspective of the study was principally the employer (public health service), and therefore the state. Since public laboratories in Australia are state-operated, we did not consider the national (Australian Commonwealth) perspective.

### Time Horizon and Indexation

In this current study, we evaluated the costs in the base year (2023-24). We evaluated these in the context of current value since these costs occurred within the relevant financial year. They are reported in 2023-24 Australian Dollars (AUD).

### Rationale and description of the model

The model utilises a deterministic range based on the upper and lower limits of the base salary reported in the industrial instruments. Inputs were directly collected where available, or an expert estimate was made. This base cost was then layered with the relevant employment conditions and entitlements (e.g. leave loading). We created an initial single composite estimate for each workforce type, which was a simple mean of the mid- point of each workforce cost range, bounded by the minimum and maximum values across the states for each labour type. We subsequently iterated this to a weighted mean, to proportionally reflect the relative costs from the contributing states with the aim of better reflecting an overall cost for Australia. The weightings were based on the Medicare enrolments, as of December 31, 2023 ^14^.

### Analytics and assumptions

A direct comparison of estimated costs and relative costs was undertaken, both overall and disaggregated. Assumptions were made based on practices in the author’s employing institution, a public health organisation in South Australia. Rights of Private practice (RoPP) were assumed to have been foregone for medical practitioners. The model assumes that individuals stay at one pay level in a given financial year, that there are no changes in base salary or components in any given year, and that many other aspects of the cost of labour remain static (such as sick leave and penalty levels). Finally, the model assumes that all allowances are completely taken up (such as professional development allowances).

### Characterising heterogeneity and uncertainty

Costs were analysed by jurisdiction and employee type, as well as creating a generalised estimate (for all jurisdictions, and thereby for Australia as a whole) and the components of labour costs were presented in disaggregated form. Deterministic Sensitivity Analysis (DSA; Scenario Analysis) was undertaken around all input parameters, including the amount of leave taken, the superannuation contribution, entitlements (e.g. continuing professional development allowance), and on the level of superannuation paid. For these, the inputs were varied stepwise. The base salary was varied based on the inputs extracted from the industrial instruments, from there all other components were varied based either on the full range of inputs collected across all jurisdictions or based on an expert estimate of the range (e.g. sick leave utilisation). The analysis of the superannuation level is based on the interaction between state and Commonwealth policy around superannuation policy settings ^15^.

Probabilistic sensitivity analysis was undertaken by fitting a gamma distribution to the range estimates to create a prior with the method of moments and then optimising using the maximum likelihood estimation method with the SciKit-Learn, Pandas and NumPy libraries ^16–18^. Monte Carlo simulation was undertaken using 10,000 samples from each distribution, and results were visualised using a histogram and kernel density estimation, with the Matplotlib and Seaborn libraries ^19, 20^. Confidence intervals were estimated using bootstrapping.

## Results

### Labour type and components

Process mapping identified five different labour types, Laboratory Technician (LT), Medical Scientist (MS), Administrative Staff (AS), Bioinformatics staff (BI) and Medical Practitioner (MP). Twelve components were identified as inputs of the cost of labour modelling: superannuation rate, working hours per week, penalties (such as overtime or weekend penalties), public holiday penalties, private practice loading, attraction and retention allowances, sick leave, professional development allowance, annual leave entitlement, annual leave loading, and base salary. Labour model inputs are detailed in Supplemental Table 1.

### Estimates of cost by labour type

Estimates of base case unit cost (cost per hour) for these clinical genomic testing labour types are summarised in Table 1 and detailed by state in Table 2. The estimated weighted mean cost per hour (minimum and maximum values) for laboratory technicians was AUD 99 (AUD 62 - AUD 142), for medical scientists was AUD 134 (AUD 80 - AUD 186), for admin staff was AUD 81 (AUD 55 - AUD 118), for bioinformatics staff was AUD 121 (AUD 80 - AUD 178), and for medical staff was AUD 368 (AUD 263 - AUD 467).

**Table 1.** Overall Estimates of Public Health Clinical Genomics Laboratory Labour, by Labour Type, 2023-24 Australian Dollars (AUD $)

| Personnel type | Mean Labour Cost (Range), Total | Mean Labour Cost (Range), Hourly |
| --- | --- | --- |
| Lab technician | 196,080 (117,643 - 281,453) | 99 (62 - 142) |
| Medical scientist | 260,925 (157,538 - 366,928) | 134 (80 - 186) |
| Admin staff | 160,794 (113,848 - 233,350) | 81 (55 - 118) |
| Bioinformatics staff | 241,018 (156,865 - 334,774) | 121 (80 - 178) |
| Pathologists | 703,206 (548,011 - 923,661) | 348 (263 - 467) |

**Table 2.** Table of Total Costs and Cost Components for Public Health Clinical Genomics Laboratory Labour, by Labour Type and Jurisdiction, 2023-24 Australian Dollars (AUD) ALL, Range and Mean from all categories; QLD, Queensland; NSW, New South Wales; SA, South Australia; WA, Western Australia; VIC, Victoria

| State | Personnel type | Labour Cost (Range), Total | Labour Cost (Range), Hourly |
| --- | --- | --- | --- |
| VIC | Lab technician | 242,302 (203,151 - 281,453) | 123 (103 - 142) |
|  | Medical scientist | 262,233 (157,538 - 366,928) | 133 (80 - 186) |
|  | Admin staff | 197,938 (162,526 - 233,350) | 100 (82 - 118) |
|  | Bioinformatics staff | 259,887 (203,151 - 316,622) | 132 (103 - 160) |
|  | Pathologists | 755,218 (640,547 - 869,888) | 382 (324 - 440) |
| SA | Lab technician | 183,535 (128,458 - 238,613) | 94 (66 - 122) |
|  | Medical scientist | 248,311 (158,142 - 338,480) | 127 (81 - 174) |
|  | Admin staff | 149,748 (134,488 - 165,008) | 77 (69 - 85) |
|  | Bioinformatics staff | 216,414 (156,865 - 275,963) | 111 (80 - 142) |
|  | Pathologists | 815,461 (707,262 - 923,661) | 413 (358 - 467) |
| NSW | Lab technician | 183,452 (141,818 - 225,086) | 88 (68 - 108) |
|  | Medical scientist | 262,537 (171,514 - 353,559) | 133 (87 - 179) |
|  | Admin staff | 136,915 (113,848 - 159,982) | 66 (55 - 77) |
|  | Bioinformatics staff | 230,733 (168,662 - 292,805) | 111 (81 - 141) |
|  | Pathologists | 637,526 (548,011 - 727,040) | 307 (263 - 350) |
| QLD | Lab technician | 169,273 (117,643 - 220,903) | 90 (62 - 117) |
|  | Medical scientist | 250,634 (166,494 - 334,774) | 133 (88 - 178) |
|  | Admin staff | 154,022 (115,620 - 192,424) | 82 (61 - 102) |
|  | Bioinformatics staff | 250,634 (166,494 - 334,774) | 133 (88 - 178) |
|  | Pathologists | 650,304 (549,076 - 751,532) | 329 (278 - 380) |
| WA | Lab technician | 184,009 (148,456 - 219,561) | 93 (75 - 111) |
|  | Medical scientist | 281,141 (200,677 - 361,605) | 142 (102 - 183) |
|  | Admin staff | 163,243 (148,456 - 178,030) | 83 (75 - 90) |
|  | Bioinformatics staff | 223,748 (206,002 - 241,495) | 113 (104 - 122) |
|  | Pathologists | 803,136 (702,710 - 903,563) | 386 (338 - 434) |

### Contribution of labour cost component to overall labour cost

Estimated labour component cost is detailed in Supplemental Table 2 and as a proportion of unit cost is detailed in Supplemental Table 3. In LT, the largest costs were attributable to base salary, at 32.3% (29.0% - 39.2%) of the total cost, followed by superannuation at 4.3% (4.1% - 5.0%) and penalties at 3.9% (3.9% - 3.9%). MS, AS and BS followed a similar pattern. For PT, base salary remained the largest component 35.3% (27.9% - 45.3%), with superannuation 4.3% (4.1% - 5.0%), following along with leave loading 4.1% (0.0% - 6.9%) and professional development 3.8% (2.8% - 5.0%). Professional development was a cost that was considerably different between MP and the other labour types.

### Sensitivity analyses

A summary of the DSAs undertaken is presented in Figure 2, Supplemental Table 4, and Supplemental Figure 2. The expected cost (mean, 95% confidence interval) for AS was 89 (88 - 89) per hour, for BS was 125 (124 - 126) per hour, for LT was 102 (101 - 102) per hour, for MS was 134 (133 - 135) per hour and PT was 378 (376 - 380) per hour. The results of the PSA are presented in Supplemental Tables 5 and 6, and Supplemental Figures 4 and 5.

**Figure 1.**
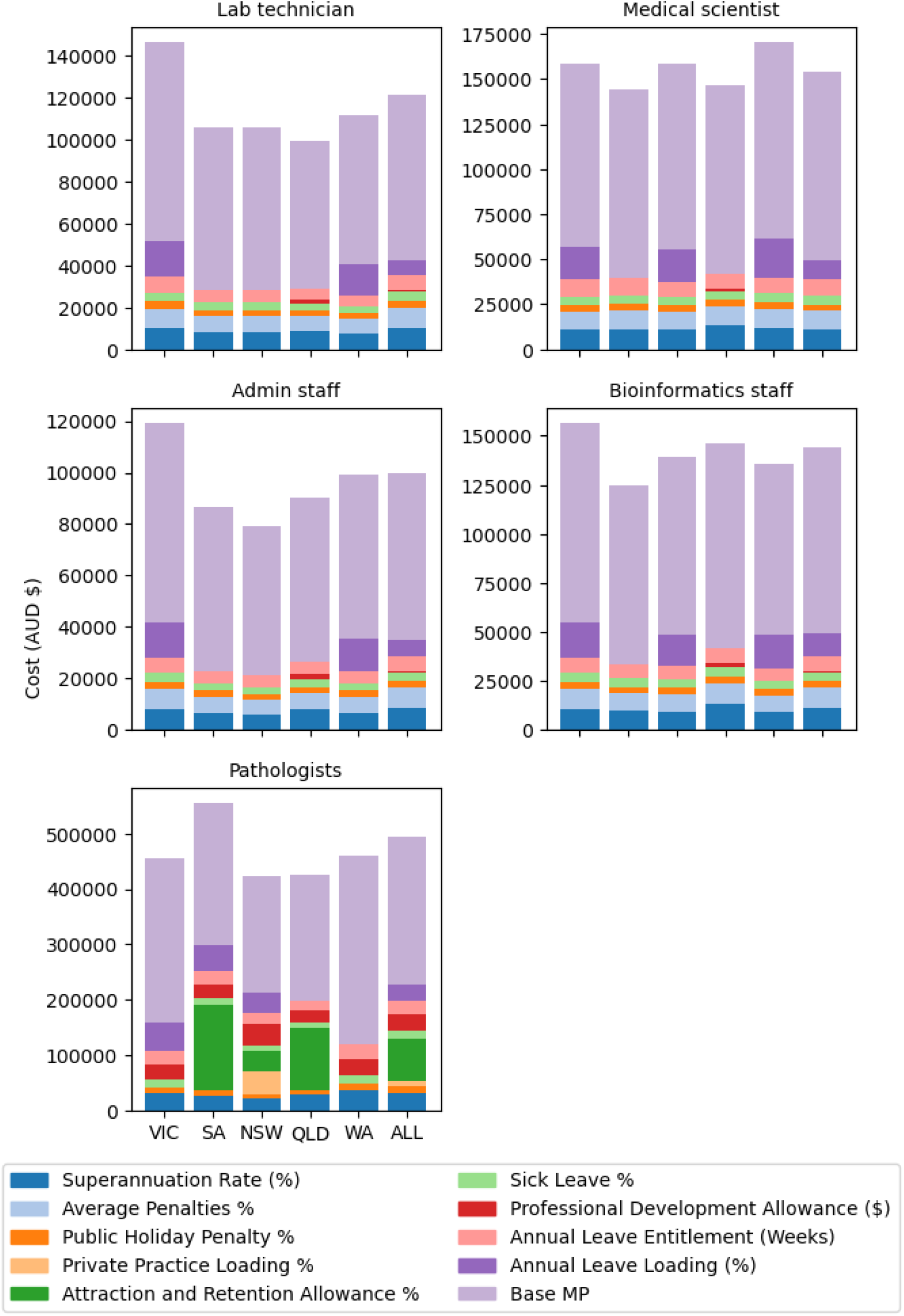
Plot of Total Costs and Cost Components for Public Health Clinical Genomics Laboratory Labour, by Labour Type and Jurisdiction, 2023-24 Australian Dollars (AUD) ALL, Range and Mean from all categories; QLD, Queensland; NSW, New South Wales; SA, South Australia; WA, Western Australia; VIC, Victoria

**Figure 2.**
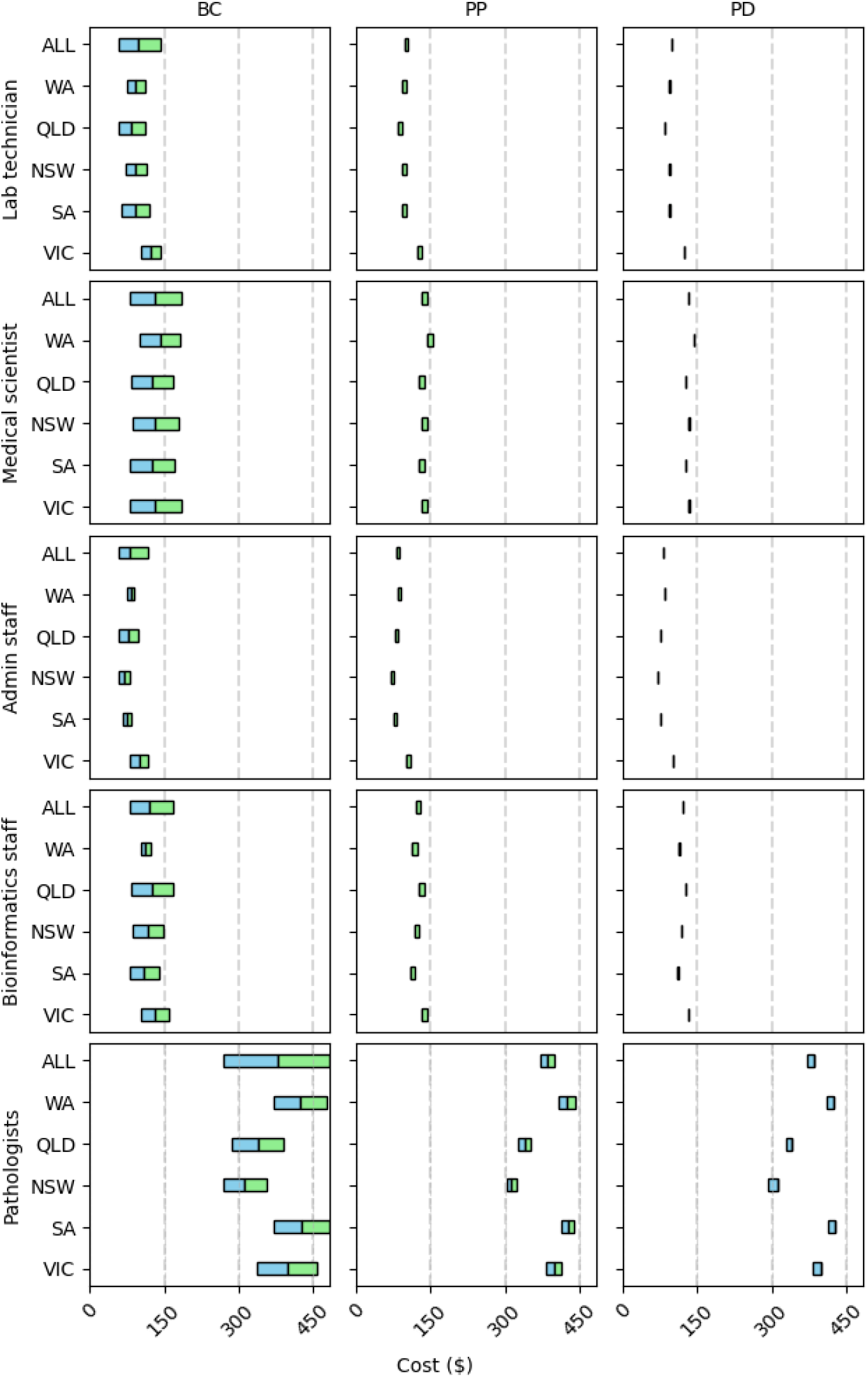
Deterministic Sensitivity Analysis of Labour Costs, by Labour Type and Jurisdiction, 2023-24 Australian Dollars (AUD) BC, Base Case; PP, Private Practice Loading; PD, Professional Development Allowance; ALL, Range and Unweighted Mean from all categories; QLD, Queensland; NSW, New South Wales; SA, South Australia; WA, Western Australia; VIC, Victoria. Tornado plot visualising the uniform distribution resulting from varying the inputs. This demonstrates the three inputs with the greatest variance, clearly demonstrating the highest variance with base salary. The values shown are estimated hourly costs based on the mean and range across states.

## Discussion

We conducted a microcosting analysis of clinical genomic testing labour workforce cost, enabling comparison within Australia across Australian jurisdictions and potentially internationally. We found that base salary accounts for the largest component of labour cost, followed by superannuation, and penalties (in all workforces except medical). In medical staff, the third highest cost component was professional development allowance. Pathologist cost was 2.9 times higher than medical scientists, 3.2 times higher than bioinformatics staff, 3.9 times higher than laboratory technicians and 4.7 times higher than the cost of administration staff. Base salary was the most significant contributor to both labour cost and cost uncertainty.

### Estimates rather than empirical

We used input estimates for some parameters that were not a part of the pay scales and associated documents, that were based on assumptions or simplifications. An example was the assumption that sick leave resulted in a mean additional cost of 6.5%. This was based on the practice within the primary author’s institution, and validated by empirical examples in the area that the primary author operationally manages.

These assumptions may not be generalisable and may be sensitive to subgroup differences. The trade-off is model simplicity vs accuracy. To assess the uncertainty around these, we undertook a DSA where we varied the sick leave percentage from 1.0-8.0%, again based on expert estimates that the range can commonly vary between this range, demonstrating that sick leave can be an important cause of cost variance.

### Generalisability

The model itself is intended to address a generalisability issue- that is to provide comparative estimates of workforce costs to enable greater understanding, translatability, and generalisability within Australia and in other countries. Built upon, this could be expanded to include other labour types, all states and territories in Australia.

### Future Research

Our study assumed that RoPP were foregone in the remuneration arrangements for medical practitioners in public health genomics laboratories. RoPP is a remuneration mechanism where certain workers (particularly medical practitioners) receive an effective performance-based component of their salary, where they share a component of private practice earnings, or undertake work that is entirely private practice. This was introduced in some states to share the cost of labour cost rises with the Australian Commonwealth. In Australian states, this is typically where individuals share a component of the Medicare Benefits Services (MBS) benefit, the Australian Commonwealth mechanism for supporting access to private practice through reimbursement. Our assumption was based on feedback during the study that this was typically foregone in laboratories (typically in exchange for a private practice allowance). Firstly, this is not necessarily the case in all public health pathology laboratories and secondly, this is not the case in non-laboratory health services. Our assumption was made on the basis that public information is not available about the amounts that individuals are paid under the RoPP arrangement. Future work could seek to estimate the amount of RoPP by professional and speciality groups. Similarly, our modelling was based on some non-empirical estimates and assumptions, such as around leave utilisation. Future work could seek to empirically measure rather than assume estimates.

We created a model useful for our specific application, however as further costing studies are undertaken, there is an opportunity to build knowledge and refine the approach to estimating workforce costs. To this end, we have provided detailed supplementary tables and the code for estimating uncertainty. We plan to build on this work with future projects, however, encourage others to expand and maintain the models, to increase the accessibility of undertaking comparative cost assessments.

### Limitations

Our study made several assumptions. Firstly, we assumed that the labour type was generalisable across Australian jurisdictions. Whilst this may be a safe assumption for medical practitioners, who are registered and regulated health professionals, there may be more variability in the other workforce types. We assumed that workforce cost was based on the public industrial agreements. In most instruments and jurisdictions there were controls identified in the instruments that limited the use of “out-of-instrument" payments, suggesting that the assumption was reasonable. Finally, there are other costs that we have not included- such as insurance. These may be context-specific and the microcosting approach lends itself to adaptation for contexts that warrant inclusion of these costs.

### Implications for Policy and Practice

The key implication of this framework and output is a reproducible way to identify bottom-up costs contributing to the labour cost of health services and health technologies, enabling comparative health research within Australia, and overseas. The specific application in this paper was clinical genomic testing, a complex health technology that is rapidly changing and in which automation is already a key consideration.

## Conclusion

We undertook microcosting of public clinical genomic testing labour costs in an Australian health services context. We identified that there are five labour types, with 12 components contributing to the estimated cost of labour. To explore the uncertainty in this modelling, we undertook scenario analyses (DSA), which identified that base salary was the greatest source of cost, as well as potential variance and uncertainty. This framework provides a reproducible way for estimated disaggregated workforce costs, that could be built upon or adapted for other economic and policy contexts

## Ethics approval

This research did not involve research on human research participants and was therefore exempt from research ethics processes in our institutions.

## Data Availability Statement

Data used in this study is provided in supplementary tables, documents and an archive of the original documents, including the publicly available source documents (pay scales, awards and enterprise bargaining agreements). These are not cited individually, as there are a large number and the documents themselves are industrial rather than academic publications.

## Data Availability

Supplementary data available

## Acknowledgements

Professor Kim Dalziel, Professor Zornitza Stark and Associate Professor Ilias Goranitis contributed to the conception and planning of the primary article on microcosting clinical genomic testing, of which the labour costing model formed a part. Professor’s Dalziel and Stark contributed to the writing and supervision of this manuscript.

