## Supplemental Document for "Standardising Workforce Cost Estimates Across Australian Jurisdictions: Genomic Testing as a Use Case"

Table 1. Table of Cost Components for Public Health Clinical Genomics Laboratory Labour, by Labour Type

| Personnel type | Lab Technician | Medical Scientist | Admin Staff | Bioinformatics Staff | Pathologists |
| --- | --- | --- | --- | --- | --- |
| Superannuation rate (%) | 11.0% | 11.0% | 11.0% | 11.0% | 11.0% |
| Hours per week | 38.0 | 37.6 | 38.0 | 38.0 | 38.8 |
| Average penalties (%) | 10.0% | 10.0% | 10.0% | 10.0% | 0.0% |
| Public holiday penalties (%) | 3.5% | 3.5% | 3.5% | 3.5% | 3.5% |
| Private practice loading (%) | 0% | 0% | 0% | 0% | 4% |
| Attraction and retention allowance (%) | 0.0% | 0.0% | 0.0% | 0.0% | 25.5% |
| Sick leave (%) | 4.6% | 4.6% | 4.6% | 4.6% | 4.6% |
| Professional development allowance ($) | $365 | $365 | $365 | $365 | $28,682 |
| Annual leave entitlement (Weeks) | 4.0 | 4.4 | 4.0 | 4.0 | 4.4 |
| Annual leave loading (%) | 7.5% | 11.0% | 7.5% | 11.0% | 10.5% |
| Indexation per annum (%) | 1.5% | 1.5% | 1.5% | 1.5% | 1.5% |
| Base salary ($) | 78,369 (48,552 - 110,314) | 104,472 (61,204 - 142,553) | 65,203 (47,704 - 91,460) | 94,994 (66,106 - 139,576) | 266,292 (179,078 - 386,515) |

Table 2. Table of Labour Components, by Labour Type and Jurisdiction, 2023-24 Australian Dollars (AUD)

AL, Annual Leave; A+R, Attraction and Retention Allowance; PL, Penalties; HW, Hours Worked; LL, Leave Loading; PP, Private Practice Loading; PD, Professional Development Allowance; PH, Public Holiday Penalties; SL, Sick Leave; SR, Superannuation; ALL, Range and Mean from all categories; QLD, Queensland; NSW, New South Wales; SA, South Australia; WA, Western Australia; VIC, Victoria

| Jurisdiction | Personnel type | SR | AP | PH | PP | A+R | SL | PD | AL | LL | Base Salary – Lower Limit | Base Salary -  Mid-Point | Base Salary -  Upper Limit |
| --- | --- | --- | --- | --- | --- | --- | --- | --- | --- | --- | --- | --- | --- |
| NSW | LT | $8,152 | $7,764 | $2,717 | $0 | $0 | $3,571 | $0 | $5,972 | $0 | $60,018 | $77,638 | $95,257 |
| NSW | MS | $10,804 | $10,290 | $3,601 | $0 | $0 | $4,733 | $0 | $7,915 | $18,007 | $67,224 | $102,900 | $138,575 |
| NSW | AS | $6,084 | $5,794 | $2,028 | $0 | $0 | $2,665 | $0 | $4,457 | $0 | $48,181 | $57,943 | $67,705 |
| NSW | BS | $9,496 | $9,043 | $3,165 | $0 | $0 | $4,160 | $0 | $6,957 | $15,826 | $66,106 | $90,435 | $114,763 |
| NSW | PT | $22,103 | $0 | $7,368 | $42,102 | $36,629 | $9,683 | $38,000 | $20,241 | $36,839 | $179,078 | $210,509 | $241,940 |
| QLD | LT | $8,950 | $7,020 | $2,457 | $0 | $0 | $3,229 | $1,826 | $5,400 | $0 | $48,552 | $70,196 | $91,840 |
| QLD | MS | $13,299 | $10,430 | $3,651 | $0 | $0 | $4,798 | $1,826 | $8,023 | $0 | $69,031 | $104,304 | $139,576 |
| QLD | AS | $8,135 | $6,380 | $2,233 | $0 | $0 | $2,935 | $1,826 | $4,908 | $0 | $47,704 | $63,803 | $79,901 |
| QLD | BS | $13,299 | $10,430 | $3,651 | $0 | $0 | $4,798 | $1,826 | $8,023 | $0 | $69,031 | $104,304 | $139,576 |
| QLD | PT | $28,783 | $0 | $7,901 | $0 | $112,874 | $10,384 | $21,500 | $17,365 | $0 | $189,406 | $225,748 | $262,090 |
| SA | LT | $8,156 | $7,767 | $2,719 | $0 | $0 | $3,573 | $0 | $5,975 | $0 | $54,364 | $77,673 | $100,982 |
| SA | MS | $10,945 | $10,424 | $3,648 | $0 | $0 | $4,795 | $0 | $10,023 | $0 | $66,386 | $104,238 | $142,090 |
| SA | AS | $6,654 | $6,337 | $2,218 | $0 | $0 | $2,915 | $0 | $4,875 | $0 | $56,916 | $63,374 | $69,832 |
| SA | BS | $9,617 | $9,159 | $3,206 | $0 | $0 | $4,213 | $0 | $7,045 | $0 | $66,386 | $91,588 | $116,789 |
| SA | PT | $26,966 | $0 | $8,989 | $0 | $154,092 | $11,814 | $26,000 | $24,694 | $44,944 | $221,622 | $256,821 | $292,019 |
| VIC | LT | $9,972 | $9,497 | $3,324 | $0 | $0 | $4,369 | $0 | $7,305 | $16,620 | $79,624 | $94,969 | $110,314 |
| VIC | MS | $10,697 | $10,188 | $3,566 | $0 | $0 | $4,686 | $0 | $9,796 | $17,829 | $61,204 | $101,878 | $142,553 |
| VIC | AS | $8,146 | $7,758 | $2,715 | $0 | $0 | $3,569 | $0 | $5,968 | $13,577 | $63,701 | $77,581 | $91,460 |
| VIC | BS | $10,695 | $10,186 | $3,565 | $0 | $0 | $4,686 | $0 | $7,835 | $17,826 | $79,624 | $101,861 | $124,098 |
| VIC | PT | $31,106 | $0 | $10,369 | $0 | $0 | $13,627 | $29,000 | $22,788 | $51,843 | $249,470 | $296,248 | $343,026 |
| WA | LT | $7,494 | $7,137 | $2,498 | $0 | $0 | $3,283 | $0 | $5,490 | $14,274 | $57,579 | $71,368 | $85,157 |
| WA | MS | $11,449 | $10,904 | $3,816 | $0 | $0 | $5,016 | $0 | $8,388 | $21,808 | $77,833 | $109,041 | $140,249 |
| WA | AS | $6,648 | $6,331 | $2,216 | $0 | $0 | $2,912 | $0 | $4,870 | $12,663 | $57,579 | $63,314 | $69,049 |
| WA | BS | $9,112 | $8,678 | $3,037 | $0 | $0 | $3,992 | $0 | $6,675 | $17,356 | $79,898 | $86,781 | $93,664 |
| WA | PT | $35,924 | $0 | $11,975 | $0 | $0 | $15,738 | $28,909 | $26,318 | $0 | $297,757 | $342,136 | $386,515 |
| ALL | LT | $10,399 (9,972 - 12,109) | $9,497 (9,497 - 9,497) | $3,324 (3,324 - 3,324) | $0 (0 - 0) | $0 (0 - 0) | $4,369 (4,369 - 4,369) | $365 (00 - 1,826) | $7,305 (7,305 - 7,305) | $7,123 (00 - 18,994) | $60,027 (48,552 - 79,624) | $78,369 (70,196 - 94,969) | $96,710 (85,157 - 110,314) |
| ALL | MS | $11,156 (10,697 - 12,989) | $10,188 (10,188 - 10,188) | $3,566 (3,566 - 3,566) | $0 (0 - 0) | $0 (0 - 0) | $4,686 (4,686 - 4,686) | $365 (00 - 1,826) | $8,620 (7,837 - 9,796) | $11,207 (00 - 20,376) | $68,336 (61,204 - 77,833) | $104,472 (101,878 - 109,041) | $140,609 (138,575 - 142,553) |
| ALL | AS | $8,495 (8,146 - 9,892) | $7,758 (7,758 - 7,758) | $2,715 (2,715 - 2,715) | $0 (0 - 0) | $0 (0 - 0) | $3,569 (3,569 - 3,569) | $365 (00 - 1,826) | $5,968 (5,968 - 5,968) | $5,819 (00 - 15,516) | $54,816 (47,704 - 63,701) | $65,203 (57,943 - 77,581) | $75,589 (67,705 - 91,460) |
| ALL | BS | $11,154 (10,695 - 12,987) | $10,186 (10,186 - 10,186) | $3,565 (3,565 - 3,565) | $0 (0 - 0) | $0 (0 - 0) | $4,686 (4,686 - 4,686) | $365 (00 - 1,826) | $7,835 (7,835 - 7,835) | $11,205 (00 - 20,372) | $72,209 (66,106 - 79,898) | $94,994 (86,781 - 104,304) | $117,778 (93,664 - 139,576) |
| ALL | PT | $32,439 (31,106 - 37,772) | $00 (00 - 00) | $10,369 (10,369 - 10,369) | $11,850 (00 - 59,250) | $75,484 (00 - 177,749) | $13,627 (13,627 - 13,627) | $28,682 (21,500 - 38,000) | $25,067 (22,788 - 28,485) | $31,106 (00 - 51,843) | $227,467 (179,078 - 297,757) | $266,292 (210,509 - 342,136) | $305,118 (241,940 - 386,515) |

Table 3. Table of Labour Components, by Labour Type and Jurisdiction, Relative Proportion (%)

AL, Annual Leave; A+R, Attraction and Retention Allowance; PL, Penalties; HW, Hours Worked; LL, Leave Loading; PP, Private Practice Loading; PD, Professional Development Allowance; PH, Public Holiday Penalties; SL, Sick Leave; SUP, Superannuation; ALL, Range and Mean from all categories; QLD, Queensland; NSW, New South Wales; SA, South Australia; WA, Western Australia; VIC, Victoria

| Jurisdiction | Personnel type | SUP | PL | PH | PP | A+R | SL | PD | AL | LL | Base Salary – Lower Limit | Base Salary -  Mid-Point | Base Salary -  Upper Limit |
| --- | --- | --- | --- | --- | --- | --- | --- | --- | --- | --- | --- | --- | --- |
| NSW | LT | 4.4% | 4.2% | 1.5% | 0.0% | 0.0% | 1.9% | 0.0% | 3.3% | 0.0% | 32.7% | 42.3% | 51.9% |
| NSW | MS | 4.1% | 3.9% | 1.4% | 0.0% | 0.0% | 1.8% | 0.0% | 3.0% | 6.9% | 25.6% | 39.2% | 52.8% |
| NSW | AS | 4.4% | 4.2% | 1.5% | 0.0% | 0.0% | 1.9% | 0.0% | 3.3% | 0.0% | 35.2% | 42.3% | 49.5% |
| NSW | BS | 4.1% | 3.9% | 1.4% | 0.0% | 0.0% | 1.8% | 0.0% | 3.0% | 6.9% | 28.7% | 39.2% | 49.7% |
| NSW | PT | 3.5% | 0.0% | 1.2% | 6.6% | 5.7% | 1.5% | 6.0% | 3.2% | 5.8% | 28.1% | 33.0% | 37.9% |
| QLD | LT | 5.3% | 4.1% | 1.5% | 0.0% | 0.0% | 1.9% | 1.1% | 3.2% | 0.0% | 28.7% | 41.5% | 54.3% |
| QLD | MS | 5.3% | 4.2% | 1.5% | 0.0% | 0.0% | 1.9% | 0.7% | 3.2% | 0.0% | 27.5% | 41.6% | 55.7% |
| QLD | AS | 5.3% | 4.1% | 1.4% | 0.0% | 0.0% | 1.9% | 1.2% | 3.2% | 0.0% | 31.0% | 41.4% | 51.9% |
| QLD | BS | 5.3% | 4.2% | 1.5% | 0.0% | 0.0% | 1.9% | 0.7% | 3.2% | 0.0% | 27.5% | 41.6% | 55.7% |
| QLD | PT | 4.4% | 0.0% | 1.2% | 0.0% | 17.4% | 1.6% | 3.3% | 2.7% | 0.0% | 29.1% | 34.7% | 40.3% |
| SA | LT | 4.4% | 4.2% | 1.5% | 0.0% | 0.0% | 1.9% | 0.0% | 3.3% | 0.0% | 29.6% | 42.3% | 55.0% |
| SA | MS | 4.4% | 4.2% | 1.5% | 0.0% | 0.0% | 1.9% | 0.0% | 4.0% | 0.0% | 26.7% | 42.0% | 57.2% |
| SA | AS | 4.4% | 4.2% | 1.5% | 0.0% | 0.0% | 1.9% | 0.0% | 3.3% | 0.0% | 38.0% | 42.3% | 46.6% |
| SA | BS | 4.4% | 4.2% | 1.5% | 0.0% | 0.0% | 1.9% | 0.0% | 3.3% | 0.0% | 30.7% | 42.3% | 54.0% |
| SA | PT | 3.3% | 0.0% | 1.1% | 0.0% | 18.9% | 1.4% | 3.2% | 3.0% | 5.5% | 27.2% | 31.5% | 35.8% |
| VIC | LT | 4.1% | 3.9% | 1.4% | 0.0% | 0.0% | 1.8% | 0.0% | 3.0% | 6.9% | 32.9% | 39.2% | 45.5% |
| VIC | MS | 4.1% | 3.9% | 1.4% | 0.0% | 0.0% | 1.8% | 0.0% | 3.7% | 6.8% | 23.3% | 38.9% | 54.4% |
| VIC | AS | 4.1% | 3.9% | 1.4% | 0.0% | 0.0% | 1.8% | 0.0% | 3.0% | 6.9% | 32.2% | 39.2% | 46.2% |
| VIC | BS | 4.1% | 3.9% | 1.4% | 0.0% | 0.0% | 1.8% | 0.0% | 3.0% | 6.9% | 30.6% | 39.2% | 47.8% |
| VIC | PT | 4.1% | 0.0% | 1.4% | 0.0% | 0.0% | 1.8% | 3.8% | 3.0% | 6.9% | 33.0% | 39.2% | 45.4% |
| WA | LT | 4.1% | 3.9% | 1.4% | 0.0% | 0.0% | 1.8% | 0.0% | 3.0% | 7.8% | 31.3% | 38.8% | 46.3% |
| WA | MS | 4.1% | 3.9% | 1.4% | 0.0% | 0.0% | 1.8% | 0.0% | 3.0% | 7.8% | 27.7% | 38.8% | 49.9% |
| WA | AS | 4.1% | 3.9% | 1.4% | 0.0% | 0.0% | 1.8% | 0.0% | 3.0% | 7.8% | 35.3% | 38.8% | 42.3% |
| WA | BS | 4.1% | 3.9% | 1.4% | 0.0% | 0.0% | 1.8% | 0.0% | 3.0% | 7.8% | 35.7% | 38.8% | 41.9% |
| WA | PT | 4.5% | 0.0% | 1.5% | 0.0% | 0.0% | 2.0% | 3.6% | 3.3% | 0.0% | 37.1% | 42.6% | 48.1% |
| ALL | LT | 4.3% (4.1% - 5.0%) | 3.9% (3.9% - 3.9%) | 1.4% (1.4% - 1.4%) | 0.0% (0.0% - 0.0%) | 0.0% (0.0% - 0.0%) | 1.8% (1.8% - 1.8%) | 0.2% (0.0% - 0.8%) | 3.0% (3.0% - 3.0%) | 2.9% (0.0% - 7.8%) | 24.8% (20.0% - 32.9%) | 32.3% (29.0% - 39.2%) | 39.9% (35.1% - 45.5%) |
| ALL | MS | 4.3% (4.1% - 5.0%) | 3.9% (3.9% - 3.9%) | 1.4% (1.4% - 1.4%) | 0.0% (0.0% - 0.0%) | 0.0% (0.0% - 0.0%) | 1.8% (1.8% - 1.8%) | 0.1% (0.0% - 0.7%) | 3.3% (3.0% - 3.7%) | 4.3% (0.0% - 7.8%) | 26.1% (23.3% - 29.7%) | 39.8% (38.9% - 41.6%) | 53.6% (52.8% - 54.4%) |
| ALL | AS | 4.3% (4.1% - 5.0%) | 3.9% (3.9% - 3.9%) | 1.4% (1.4% - 1.4%) | 0.0% (0.0% - 0.0%) | 0.0% (0.0% - 0.0%) | 1.8% (1.8% - 1.8%) | 0.2% (0.0% - 0.9%) | 3.0% (3.0% - 3.0%) | 2.9% (0.0% - 7.8%) | 27.7% (24.1% - 32.2%) | 32.9% (29.3% - 39.2%) | 38.2% (34.2% - 46.2%) |
| ALL | BS | 4.3% (4.1% - 5.0%) | 3.9% (3.9% - 3.9%) | 1.4% (1.4% - 1.4%) | 0.0% (0.0% - 0.0%) | 0.0% (0.0% - 0.0%) | 1.8% (1.8% - 1.8%) | 0.1% (0.0% - 0.7%) | 3.0% (3.0% - 3.0%) | 4.3% (0.0% - 7.8%) | 27.8% (25.4% - 30.7%) | 36.6% (33.4% - 40.1%) | 45.3% (36.0% - 53.7%) |
| ALL | PT | 4.3% (4.1% - 5.0%) | 0.0% (0.0% - 0.0%) | 1.4% (1.4% - 1.4%) | 1.6% (0.0% - 7.8%) | 10.0% (0.0% - 23.5%) | 1.8% (1.8% - 1.8%) | 3.8% (2.8% - 5.0%) | 3.3% (3.0% - 3.8%) | 4.1% (0.0% - 6.9%) | 30.1% (23.7% - 39.4%) | 35.3% (27.9% - 45.3%) | 40.4% (32.0% - 51.2%) |

Table 4. Table of Deterministic Sensitivity Analysis (DSA) of Labour Costs (Hourly), by Labour Type and Jurisdiction, 2023-24 Australian Dollars (AUD)

ALL, Range and Mean from all categories; QLD, Queensland; NSW, New South Wales; SA, South Australia; WA, Western Australia; VIC, Victoria

| Labour Type | Variable | NSW | QLD | SA | VIC | WA | ALL |
| --- | --- | --- | --- | --- | --- | --- | --- |
|  |  | Mean (95% CI) | | | | | |
| Admin staff | Annual Leave | 69 (58 - 81) | 77 (58 - 97) | 76 (68 - 84) | 100 (82 - 118) | 83 (75 - 90) | 81 (59 - 114) |
|  | Attraction and Retention | 69 (58 - 81) | 77 (58 - 97) | 76 (68 - 84) | 100 (82 - 118) | 83 (75 - 90) | 81 (59 - 114) |
|  | Average Penalties | 69 (55 - 88) | 77 (56 - 105) | 76 (65 - 91) | 100 (79 - 128) | 83 (72 - 97) | 81 (57 - 123) |
|  | Base Salary | 69 (58 - 81) | 77 (58 - 97) | 76 (68 - 84) | 100 (82 - 118) | 83 (75 - 90) | 81 (58 - 118) |
|  | Hours worked | 69 (55 - 85) | 77 (55 - 101) | 76 (65 - 88) | 100 (78 - 124) | 83 (72 - 94) | 81 (56 - 119) |
|  | Leave Loading | 69 (58 - 81) | 77 (58 - 97) | 76 (68 - 84) | 100 (82 - 118) | 83 (75 - 90) | 81 (59 - 114) |
|  | Private Practice Loading | 69 (58 - 81) | 77 (58 - 97) | 76 (68 - 84) | 100 (82 - 118) | 83 (75 - 90) | 81 (59 - 114) |
|  | Professional Development | 69 (58 - 81) | 77 (58 - 97) | 76 (68 - 84) | 100 (82 - 118) | 83 (75 - 90) | 81 (59 - 114) |
|  | Public Holiday Penalty | 69 (58 - 81) | 77 (58 - 97) | 76 (68 - 84) | 100 (82 - 119) | 83 (75 - 90) | 81 (59 - 114) |
|  | Sick Leave | 69 (58 - 81) | 77 (58 - 97) | 76 (68 - 84) | 100 (82 - 118) | 83 (75 - 90) | 81 (59 - 114) |
|  | Superannuation | 69 (58 - 82) | 77 (58 - 97) | 76 (68 - 84) | 100 (82 - 119) | 83 (75 - 91) | 81 (59 - 114) |
| Bioinformatics staff | Annual Leave | 117 (86 - 148) | 126 (84 - 168) | 110 (80 - 140) | 132 (103 - 161) | 113 (104 - 122) | 120 (83 - 176) |
|  | Attraction and Retention | 117 (86 - 148) | 126 (84 - 168) | 110 (80 - 140) | 132 (103 - 161) | 113 (104 - 122) | 120 (83 - 176) |
|  | Average Penalties | 117 (82 - 160) | 126 (80 - 182) | 110 (76 - 152) | 132 (99 - 173) | 113 (100 - 132) | 120 (80 - 190) |
|  | Base Salary | 117 (86 - 148) | 126 (84 - 168) | 110 (80 - 140) | 132 (103 - 161) | 113 (104 - 122) | 120 (80 - 168) |
|  | Hours worked | 117 (81 - 155) | 126 (79 - 176) | 110 (76 - 147) | 132 (98 - 168) | 113 (99 - 128) | 120 (79 - 184) |
|  | Leave Loading | 117 (86 - 148) | 126 (84 - 168) | 110 (80 - 140) | 132 (103 - 161) | 113 (104 - 122) | 120 (83 - 176) |
|  | Private Practice Loading | 117 (86 - 148) | 126 (84 - 168) | 110 (80 - 140) | 132 (103 - 161) | 113 (104 - 122) | 120 (83 - 176) |
|  | Professional Development | 117 (86 - 148) | 126 (84 - 168) | 110 (80 - 140) | 132 (103 - 161) | 113 (104 - 122) | 120 (83 - 176) |
|  | Public Holiday Penalty | 117 (85 - 149) | 126 (83 - 169) | 110 (79 - 140) | 132 (103 - 161) | 113 (104 - 123) | 120 (83 - 176) |
|  | Sick Leave | 117 (86 - 148) | 126 (84 - 168) | 110 (80 - 140) | 132 (103 - 161) | 113 (104 - 122) | 120 (83 - 176) |
|  | Superannuation | 117 (85 - 149) | 126 (83 - 169) | 110 (79 - 141) | 132 (103 - 162) | 113 (104 - 123) | 120 (83 - 177) |
| Lab technician | Annual Leave | 93 (72 - 114) | 85 (59 - 111) | 93 (65 - 121) | 123 (103 - 143) | 93 (75 - 111) | 97 (60 - 137) |
|  | Attraction and Retention | 93 (72 - 114) | 85 (59 - 111) | 93 (65 - 121) | 123 (103 - 143) | 93 (75 - 111) | 97 (60 - 137) |
|  | Average Penalties | 93 (69 - 124) | 85 (57 - 120) | 93 (62 - 131) | 123 (99 - 154) | 93 (72 - 120) | 97 (58 - 148) |
|  | Base Salary | 93 (72 - 114) | 85 (59 - 111) | 93 (65 - 121) | 123 (103 - 143) | 93 (75 - 111) | 97 (59 - 143) |
|  | Hours worked | 93 (68 - 119) | 85 (56 - 116) | 93 (62 - 127) | 123 (98 - 149) | 93 (72 - 117) | 97 (57 - 143) |
|  | Leave Loading | 93 (72 - 114) | 85 (59 - 111) | 93 (65 - 121) | 123 (103 - 143) | 93 (75 - 111) | 97 (60 - 137) |
|  | Private Practice Loading | 93 (72 - 114) | 85 (59 - 111) | 93 (65 - 121) | 123 (103 - 143) | 93 (75 - 111) | 97 (60 - 137) |
|  | Professional Development | 93 (72 - 114) | 85 (59 - 111) | 93 (65 - 121) | 123 (103 - 143) | 93 (75 - 111) | 97 (60 - 137) |
|  | Public Holiday Penalty | 93 (72 - 114) | 85 (59 - 111) | 93 (65 - 121) | 123 (103 - 143) | 93 (75 - 112) | 97 (60 - 137) |
|  | Sick Leave | 93 (72 - 114) | 85 (59 - 111) | 93 (65 - 121) | 123 (103 - 143) | 93 (75 - 111) | 97 (60 - 137) |
|  | Superannuation | 93 (72 - 115) | 85 (59 - 112) | 93 (65 - 122) | 123 (103 - 144) | 93 (75 - 112) | 97 (60 - 138) |
| Medical scientist | Annual Leave | 133 (87 - 179) | 126 (84 - 168) | 126 (80 - 172) | 133 (80 - 186) | 143 (102 - 183) | 132 (77 - 180) |
|  | Attraction and Retention | 133 (87 - 179) | 126 (84 - 168) | 126 (80 - 172) | 133 (80 - 186) | 143 (102 - 183) | 132 (77 - 180) |
|  | Average Penalties | 133 (84 - 193) | 126 (80 - 182) | 126 (77 - 186) | 133 (77 - 200) | 143 (98 - 198) | 132 (74 - 195) |
|  | Base Salary | 133 (87 - 179) | 126 (84 - 168) | 126 (80 - 172) | 133 (80 - 186) | 143 (102 - 183) | 132 (80 - 186) |
|  | Hours worked | 133 (83 - 188) | 126 (79 - 176) | 126 (76 - 180) | 133 (76 - 195) | 143 (97 - 192) | 132 (74 - 189) |
|  | Leave Loading | 133 (87 - 179) | 126 (84 - 168) | 126 (80 - 172) | 133 (80 - 186) | 143 (102 - 183) | 132 (77 - 180) |
|  | Private Practice Loading | 133 (87 - 179) | 126 (84 - 168) | 126 (80 - 172) | 133 (80 - 186) | 143 (102 - 183) | 132 (77 - 180) |
|  | Professional Development | 133 (87 - 179) | 126 (84 - 168) | 126 (80 - 172) | 133 (80 - 186) | 143 (102 - 183) | 132 (77 - 180) |
|  | Public Holiday Penalty | 133 (87 - 180) | 126 (83 - 169) | 126 (80 - 172) | 133 (80 - 186) | 143 (102 - 184) | 132 (77 - 181) |
|  | Sick Leave | 133 (87 - 179) | 126 (84 - 168) | 126 (80 - 172) | 133 (80 - 186) | 143 (102 - 183) | 132 (77 - 180) |
|  | Superannuation | 133 (87 - 181) | 126 (83 - 169) | 126 (80 - 173) | 133 (80 - 187) | 143 (102 - 185) | 132 (77 - 181) |
| Pathologists | Annual Leave | 313 (269 - 356) | 339 (286 - 391) | 426 (370 - 483) | 398 (337 - 458) | 425 (371 - 478) | 384 (263 - 551) |
|  | Attraction and Retention | 313 (269 - 356) | 339 (286 - 391) | 426 (370 - 483) | 398 (337 - 458) | 425 (371 - 478) | 384 (263 - 551) |
|  | Average Penalties | 313 (269 - 393) | 339 (286 - 431) | 426 (370 - 527) | 398 (337 - 511) | 425 (371 - 536) | 384 (263 - 610) |
|  | Base Salary | 313 (269 - 356) | 339 (286 - 391) | 426 (370 - 483) | 398 (337 - 458) | 425 (371 - 478) | 380 (269 - 483) |
|  | Hours worked | 313 (255 - 373) | 339 (271 - 410) | 426 (351 - 506) | 398 (321 - 480) | 425 (353 - 500) | 384 (250 - 577) |
|  | Leave Loading | 313 (269 - 356) | 339 (286 - 391) | 426 (370 - 483) | 398 (337 - 458) | 425 (371 - 478) | 384 (263 - 551) |
|  | Private Practice Loading | 313 (269 - 356) | 339 (286 - 391) | 426 (370 - 483) | 398 (337 - 458) | 425 (371 - 478) | 384 (263 - 551) |
|  | Professional Development | 313 (269 - 356) | 339 (286 - 391) | 426 (370 - 483) | 398 (337 - 458) | 425 (371 - 478) | 384 (263 - 551) |
|  | Public Holiday Penalty | 313 (268 - 357) | 339 (285 - 392) | 426 (369 - 484) | 398 (337 - 459) | 425 (371 - 479) | 384 (263 - 552) |
|  | Sick Leave | 313 (269 - 356) | 339 (286 - 391) | 426 (370 - 483) | 398 (337 - 458) | 425 (371 - 478) | 384 (263 - 551) |
|  | Superannuation | 313 (268 - 358) | 339 (285 - 394) | 426 (369 - 486) | 398 (337 - 461) | 425 (371 - 481) | 384 (263 - 555) |

Table 5. Table of Probabilistic Sensitivity Analysis (PSA) of Labour Costs (Total), by Labour Type and Jurisdiction, 2023-24 Australian Dollars (AUD)

ALL, Range and Mean from all categories; QLD, Queensland; NSW, New South Wales; SA, South Australia; WA, Western Australia; VIC, Victoria

| Personnel type | Jurisdiction | | | | | |
| --- | --- | --- | --- | --- | --- | --- |
|  | NSW | QLD | SA | VIC | WA | ALL |
| Admin Staff | 137,620 (137,174 - 138,084) | 153,635 (152,905 - 154,357) | 150,357 (150,068 - 150,653) | 198,979 (198,312 - 199,638) | 163,834 (163,564 - 164,131) | 175,055 (173,831 - 176,261) |
| Bioinformatics Staff | 232,387 (231,167 - 233,497) | 250,307 (248,614 - 251,855) | 217,935 (216,728 - 219,090) | 261,431 (260,327 - 262,546) | 224,531 (224,187 - 224,896) | 246,137 (244,396 - 247,944) |
| Lab Technician | 184,570 (183,733 - 185,423) | 169,065 (168,018 - 170,080) | 184,877 (183,750 - 185,972) | 243,448 (242,695 - 244,168) | 185,024 (184,331 - 185,723) | 200,833 (199,111 - 202,460) |
| Medical Scientist | 264,693 (262,869 - 266,509) | 250,307 (248,614 - 251,855) | 250,242 (248,360 - 251,994) | 264,030 (261,856 - 266,218) | 283,037 (281,397 - 284,636) | 264,030 (261,856 - 266,218) |
| Pathologists | 619,023 (617,376 - 620,735) | 670,785 (668,841 - 672,714) | 844,032 (841,863 - 846,224) | 788,214 (785,987 - 790,421) | 840,397 (838,380 - 842,442) | 746,592 (742,388 - 750,591) |

Table 6. Table of Probabilistic Sensitivity Analysis (PSA) of Labour Costs (Hourly), by Labour Type and Jurisdiction, 2023-24 Australian Dollars (AUD)

ALL, Range and Mean from all categories; QLD, Queensland; NSW, New South Wales; SA, South Australia; WA, Western Australia; VIC, Victoria

| Personnel type | Jurisdiction | | | | | |
| --- | --- | --- | --- | --- | --- | --- |
|  | NSW | QLD | SA | VIC | WA | ALL |
| Admin staff | 70 (69 - 70) | 78 (77 - 78) | 76 (76 - 76) | 101 (100 - 101) | 83 (83 - 83) | 89 (88 - 89) |
| Bioinformatics staff | 118 (117 - 118) | 127 (126 - 127) | 110 (110 - 111) | 132 (132 - 133) | 114 (113 - 114) | 125 (124 - 126) |
| Lab technician | 93 (93 - 94) | 86 (85 - 86) | 94 (93 - 94) | 123 (123 - 124) | 94 (93 - 94) | 102 (101 - 102) |
| Medical scientist | 134 (133 - 135) | 127 (126 - 127) | 127 (126 - 128) | 134 (133 - 135) | 143 (143 - 144) | 134 (133 - 135) |
| Pathologists | 313 (312 - 314) | 340 (338 - 340) | 427 (426 - 428) | 399 (398 - 400) | 426 (425 - 427) | 378 (376 - 380) |

Figure 1. Plot of Total Costs and Cost Components for Public Health Clinical Genomics Laboratory Labour, by Labour Type and Jurisdiction, Relative (%)

ALL, Range and Mean from all categories; QLD, Queensland; NSW, New South Wales; SA, South Australia; WA, Western Australia; VIC, Victoria

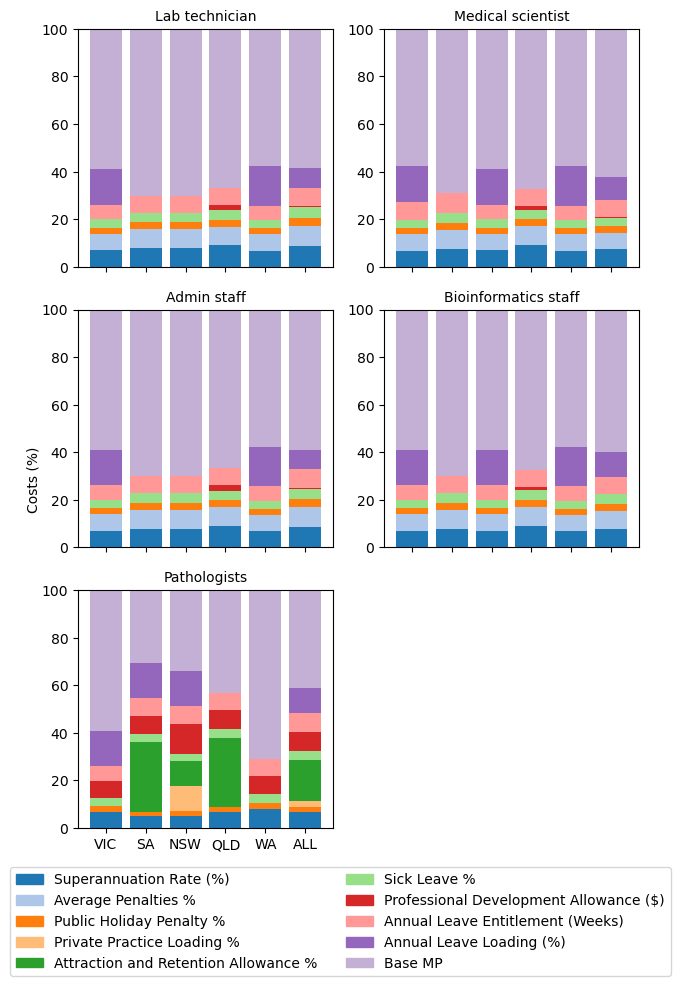

Figure 2. Deterministic Sensitivity Analysis of Labour Costs, by Labour Type and Jurisdiction, 2023-24 Australian Dollars (AUD)

AL, Annual Leave; A+R, Attraction and Retention Allowance; PL, Penalties; HW, Hours Worked; LL, Leave Loading; PP, Private Practice Loading; PD, Professional Development Allowance; PH, Public Holiday Penalties; SL, Sick Leave; SUP, Superannuation; ALL, Range and Mean from all categories; QLD, Queensland; NSW, New South Wales; SA, South Australia; WA, Western Australia; VIC, Victoria

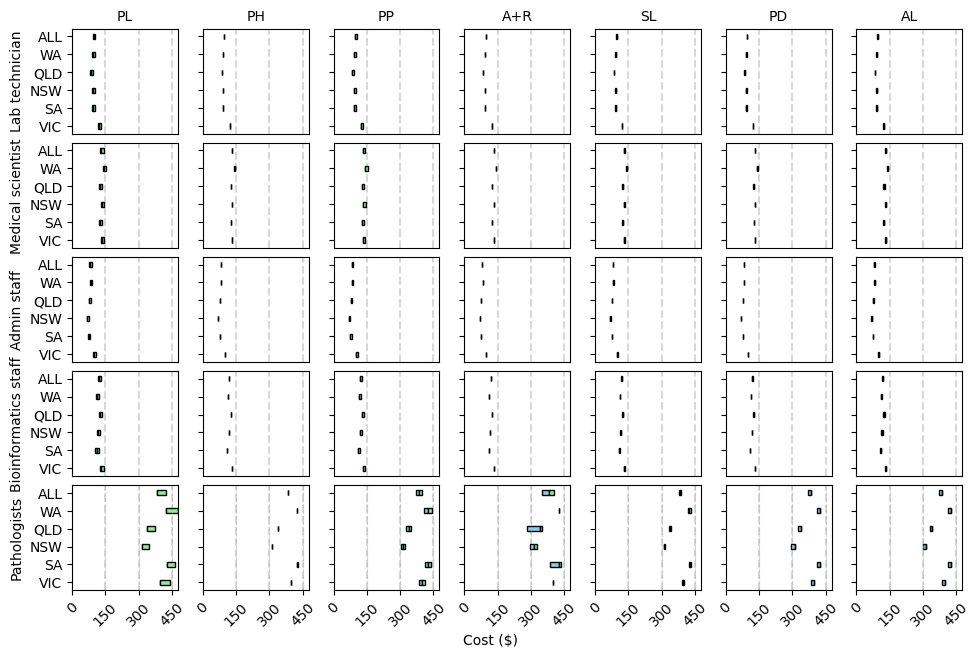

Figure 3. Probabilistic Sensitivity Analysis of Labour Costs (Total), by Labour Type and Jurisdiction, 2023-24 Australian Dollars (AUD $)

ALL, Range and Mean from all categories; QLD, Queensland; NSW, New South Wales; SA, South Australia; WA, Western Australia; VIC, Victoria

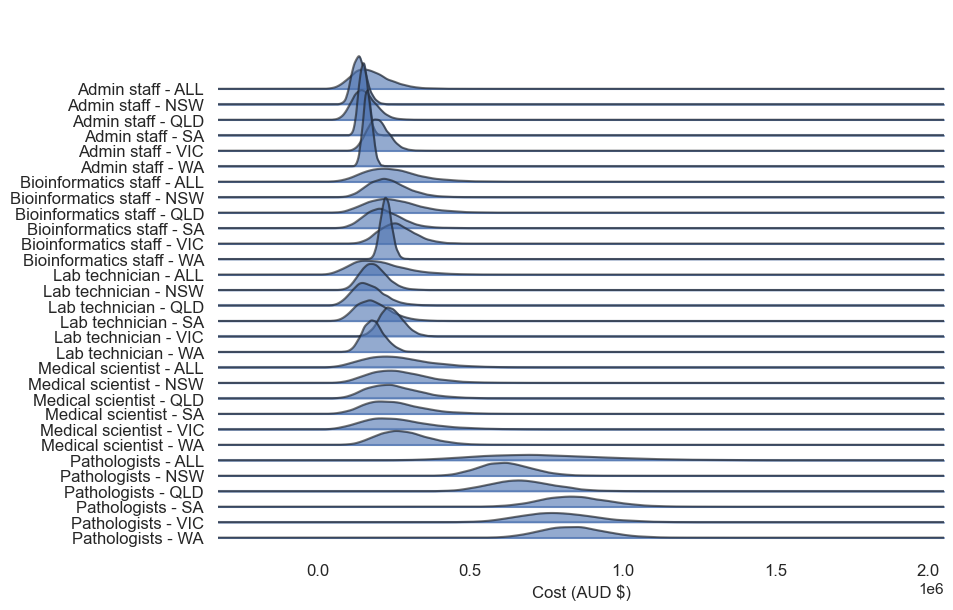

Figure 4. Probabilistic Sensitivity Analysis of Labour Costs (Hourly), by Labour Type and Jurisdiction, 2023-24 Australian Dollars (AUD $)

ALL, Range and Mean from all categories; QLD, Queensland; NSW, New South Wales; SA, South Australia; WA, Western Australia; VIC, Victoria

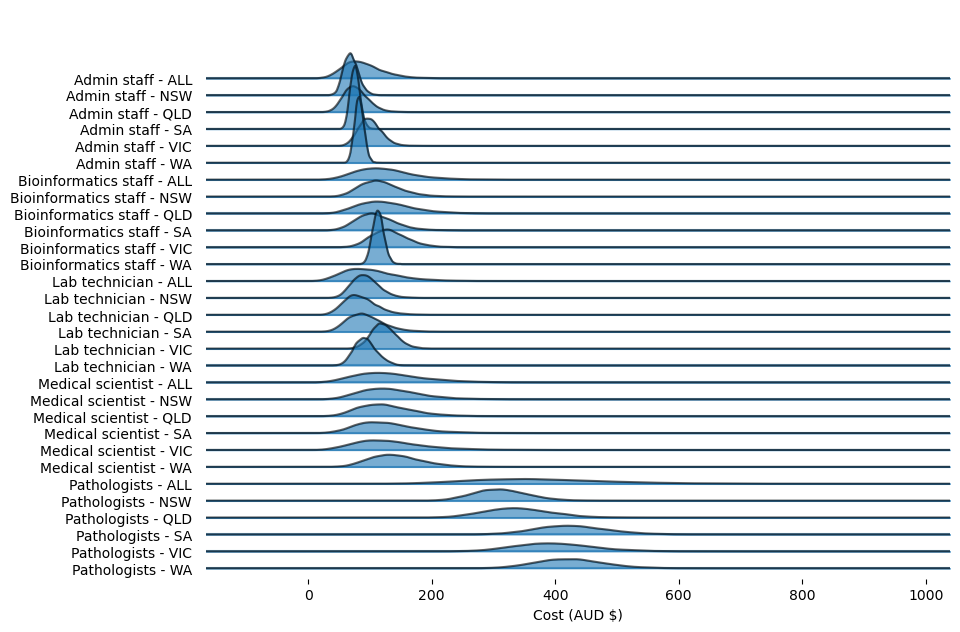
